# Predicting pathological complete response to neoadjuvant chemotherapy in breast cancer from routine diagnostic histopathology biopsies

**DOI:** 10.1101/2022.11.11.22282205

**Authors:** Witali Aswolinskiy, Enrico Munari, Hugo M. Horlings, Lennart Mulder, Giuseppe Bogina, Joyce Sanders, Yat-Hee Liu, Alexandra W. van den Belt-Dusebout, Leslie Tessier, Maschenka Balkenhol, Jeffrey Hoven, Jelle Wesseling, Jeroen van der Laak, Esther H. Lips, Francesco Ciompi

**Author notes:** equal contribution to the work (Enrico Munari, Hugo M. Horlings, Esther H. Lips, Francesco Ciompi).

## Abstract

**Purpose:** Invasive breast cancer patients are increasingly being treated with neoadjuvant chemotherapy, however, only a fraction of the patients respond to it completely. To prevent over-treating patients with a toxic drug, there is an urgent need for biomarkers capable of predicting treatment response before administering the therapy. In this retrospective study, we developed interpretable, deep-learning based biomarkers to predict the pathological complete response (pCR, i.e. the absence of tumor cells in the surgical resection specimens) to neoadjuvant chemotherapy from digital pathology H&E images of pre-treatment breast biopsies.

**Experimental Design:** Our approach consists of two steps: In the first step, using deep learning, mitoses are detected and the tissue segmented into several morphology compartments including tumor, lymphocytes and stroma. In the second step, computational biomarkers are derived from the segmentation and detection output to encode slide-level relationships between the morphological structures with focus on tumor infiltrating lymphocytes (TILs). We developed and evaluated our method on slides from N=721 patients from three European medical centers with triple-negative and Luminal B breast cancers.

**Results:** The investigated biomarkers yield statistically significant prediction performance for pCR with areas under the receiver operating characteristic curve between 0.66 and 0.88 depending on the cancer subtype and center.

**Conclusion:** The proposed computational biomarkers predict pathological complete response, but will require more evaluation and finetuning for clinical application. The results further corroborate the potential role of deep learning to automate TILs quantification, and their predictive value in breast cancer neoadjuvant treatment planning.

## 1. INTRODUCTION

### Breast cancer and neoadjuvant chemotherap

Invasive breast cancer (IBC) is increasingly being treated with chemotherapy administered *prior* to breast cancer surgery [1]. This neoadjuvant chemotherapy (NAC) is intended to reduce the tumor load and may result in the pathological complete response (pCR), i.e., the absence of visible tumor cells in the surgery resections. Studies have shown that pCR is associated with event-free survival and recurrence-free survival [2]. However, only a fraction of treated patients responds to the treatment, with response rates that vary with the molecular subtypes of breast cancer. About 40% of patients with triple-negative breast cancers (TNBC) will achieve pCR, whereas the response rate for Luminal B breast cancer patients is only about 15% [3, 4].

Administering NAC is a process that lasts for several weeks, has side effects and de-facto postpones the surgery while the tumor may progress locally and systemically if the patient does not respond to NAC. This shows the urgent need for predicting whether treating a patient with NAC will result in pCR, to optimally plan the treatment strategy.

### On the prediction of NAC response from histopatholog

Several studies have shown the correlation between stromal tumor-infiltrating lymphocytes (TILs) and favorable NAC response and survival outcomes, where TILs are often quantified on hematoxylin and eosin (H&E) stained slides following the recommendations from the International TILs Working Group [5, 6, 7, 8]. The proposed visual procedure relies on the subjective identification of the region of the slide where an invasive tumor is present (so-called ”tumor bulk”), estimating the percentage of lymphocytes within the tumor-associated stroma in the bulk, and calculating a “TILs score” by averaging such percentages over multiple regions. Although effective, this visual procedure is hampered by potential ”pitfalls” such as the presence of e.g. ischemic tumor cells, small tumor nuclei, and fixation artifacts [9], and requires the mental exclusion of regions such as benign tissue and in-situ lesions.

Another biomarker that has been shown to carry predictive value for NAC response is the tumor proliferation score [10], assessed based on Ki67 immunohistochemistry staining as the percentage of tumor cells with positive nuclear staining [11, 12]. However, Ki67 staining may not be routinely available, and introduces additional costs compared to standard diagnostic H&E staining.

### Deep learning for computational biomarkers

To automate biomarker quantification, in recent years, researchers have started focusing on Deep Learning with Convolutional Neural Networks (CNN) to learn directly from images [13, 14]. Applied to digital pathology, Deep Learning can achieve the performance of trained pathologists for tasks such as tumor detection and grading, and is on the verge of being adopted into clinical practice [14]. The two most common tasks here are cell detection (predicting location and type of cells in the image), and tissue segmentation (delineating regions containing different tissue subtypes).

In the context of deep learning for computational biomarkers such as TIL scoring in breast cancer, several approaches have been recently proposed based on CNNs [15, 16, 17, 18, 19]. In most cases, these approaches segment the tissue into tumor, stroma and lymphocyte compartments and compute the TIL-score based on the compartment ratios. For example, in [18], the calculated stromal TIL density was defined as the ratio of the detected lymphocytes to the tumor-associated stroma. The authors showed that this biomarker was strongly correlated with the visual stromal TIL score and stratified the patients in the study significantly into two distinct survival groups in a cohort of 257 TNBC cases. These works, however, were evaluated only on surgery resections, not diagnostic biopsies, and did not investigate the predictive value for NAC response.

For pCR prediction, three studies have been carried out with a focus on the tumor-epithelium in H&E stained biopsies. In [20], a ’pCR score’ was directly computed from tumor-epithelium patches and evaluated on a set of 107 cases. In [21], nuclei in tumor regions were detected and multiple graph- and wavelet-based features computed, with an evaluation set of 38 cases. In [22], a multi-modal network learned from combined serial H&E and immunohistochemically stained slides and was evaluated on 30 TNBC cases. While these studies showed promising results, their evaluation was very limited or pooled several IBC subtypes together, making a stratified analysis and clinical interpretation difficult. Furthermore, these approaches lack morphological interpretability in contrast to engineered biomarkers such as TILs.

Here, we focus therefore on investigating directly interpretable computational biomarkers based on relations of different tissue morphologies. Motivated by clinical needs on breast cancer treatment, we focus our study on two specific breast cancer subtypes, namely triple-negative (TNBC: HR-,HER2-) and Luminal B (HR+,HER2-,grade 2/3) invasive breast cancers. We define here the pathological complete response to NAC as the absence of invasive cancer in the breast only (ypT0/is [23]). In this context, to the best of our knowledge, our work is the first to investigate computational biomarkers related to morphological characteristics with a focus on TILs and mitotic activity on diagnostic H&E-stained biopsies from multiple centers stratified in different breast cancer subtypes. Furthermore, we compare the predictive performance for pCR of the computational biomarkers and the visually assessed TILs.

## 2. METHODS

Our approach consists of two parts, visualized in Fig. 1. First, we trained a CNN to segment the slides into the classes tumor, stroma, lymphocytes, necrosis, fat and rest. We also used an existing CNN model for mitosis detection, previously validated in clinical studies [24, 25]. The output of this deep learning pipeline for a slide is a segmentation mask for the six classes and the coordinates of detected mitoses in the tumor regions. Second, we derived biomarkers from the tissue segmentation and mitoses detections, and assessed their predictive value for pCR. In this section, we introduce the used datasets and the developed methods.

**Figure 1:**
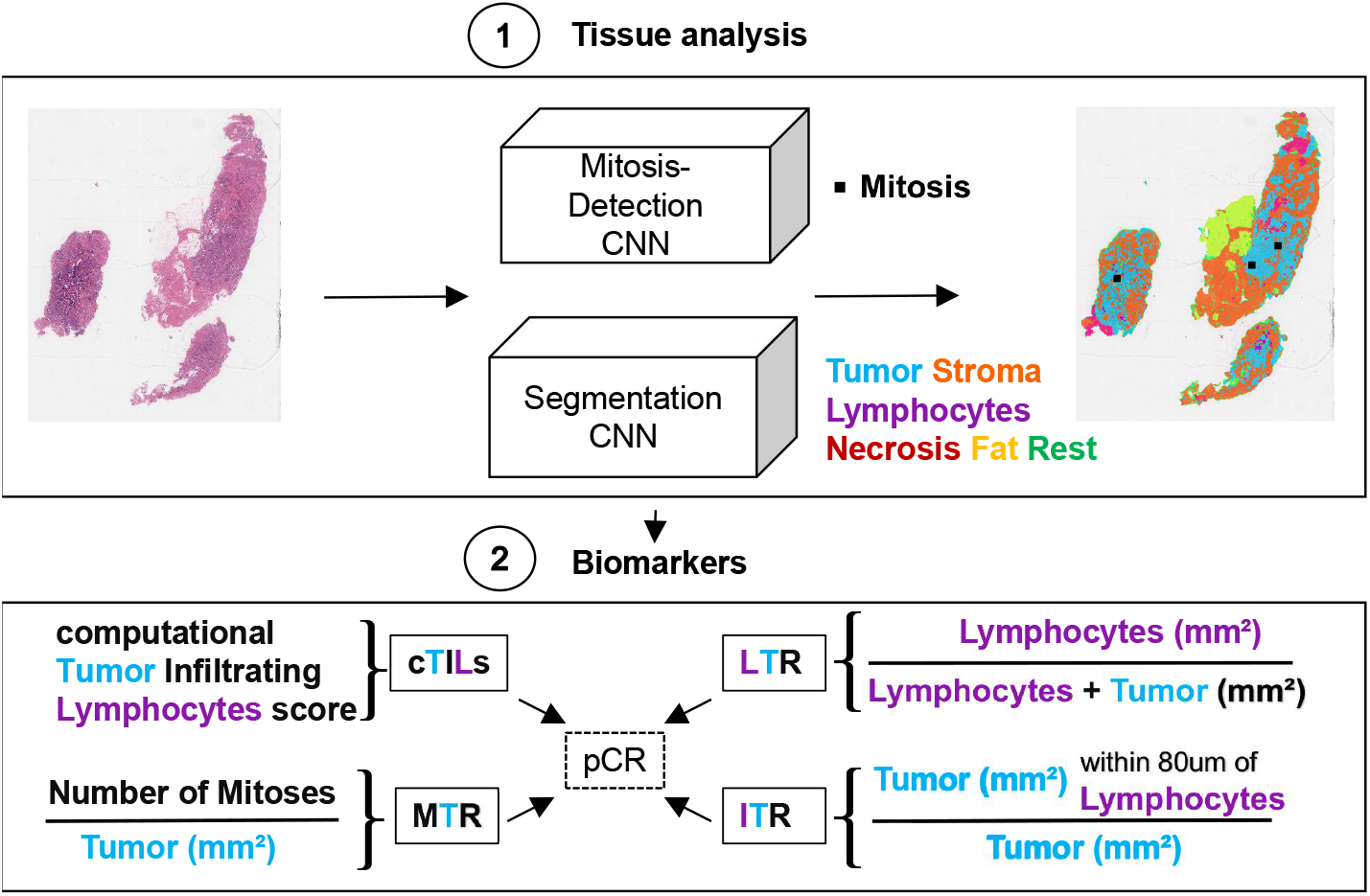
Method overview: 1) Segment slides into different tissue types and detect mitoses. 2) Compute biomarkers from the segmentation prediction of tumor, stroma and lymphocytes and detected mitoses within tumor regions. LTR: lymphocyte-tumor ratio, cTILs: computational tumor infiltrating lymphocytes score, ITR: inflamed tumor ratio (proportion of tumor close to lymphocytes), MTR: mitoses-tumor ratio.

### 2.1 Materials

#### Data for biomarker development and evaluation

Initially, we collected 911 cases from three centers: 741 from the Netherlands Cancer Institute (NKI, Amsterdam, the Netherlands), 108 from the Radboud University Medical Center (RUMC, Nijmegen, the Netherlands) and 62 from the IRCCS Sacro Cuore Don Calabria Hospital (SCDC, Verona, Italy). The NKI TNBC slides were scanned with an Aperio AT2 (Leica Biosystems) at 40X, the NKI Luminal B slides with a PANNORAMIC 1000 (3DHISTECH) scanner at 40X, the RUMC slides with a 3DHistech Pannoramic 1000 scanner at 40X and the slides from SCDC with a Ventana DP 200 slide scanner at 20X magnification. All slides are diagnostic biopsies stained with H&E, extracted via core-needle procedure (before chemotherapy). For NKI TNBC and the RUMC cases, multiple slides per case are available while the other cohorts have only one slide per case.

The slides from NKI were obtained from retrospective studies and include old glass slides. Therefore, after digitization, slides were visually inspected by pathologists, who excluded 101 slides with washed-out staining or too few tumor cells. The slides from SCDC were checked by pathologists at the time of inclusion in this study, and the RUMC slides were scanned for the purpose of this study and visually checked for quality before and after scanning, resulting in no exclusion due to quality issues. Slides from 89 NKI cases were set aside to train a deep learning model for tissue segmentation (training also included external data described in appendix section S1.2). The remaining 721 cases were randomly split into a biomarker development (N=352, NKI) and a biomarker evaluation set (N=369, all centers), with cases from RUMC and SCDC used exclusively for biomarker evaluation. The development set was used for biomarker design, e.g. choosing thresholds to maximize pCR prediction performance. The data split is visualized in Fig. 2. For all cohorts, information about the NAC response was available; additional available clinical information is listed in appendix Tables S1 and S2.

**Figure 2:**
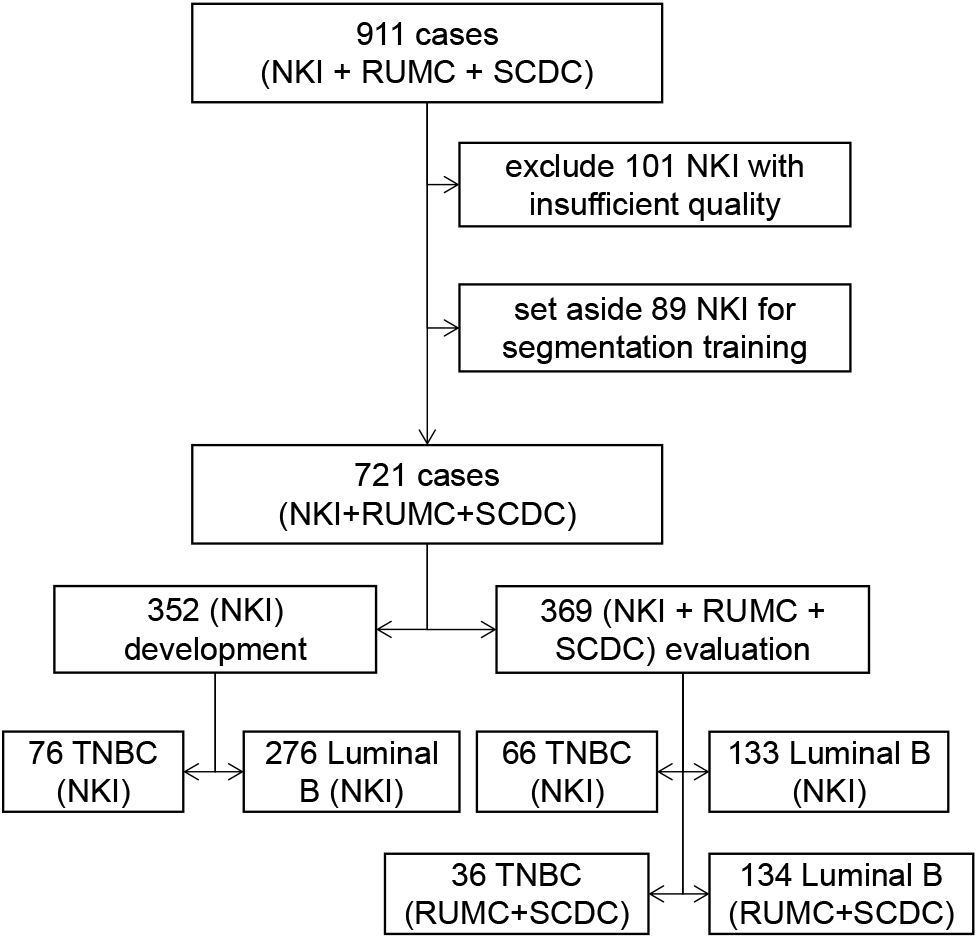
Biomarker development and validation data: visualization of the data split per type (TNBC, Luminal B), center (NKI, RUMC and SCDC) and data subset (development and evaluation), starting from the initial inclusion of cases to the definition of the development and evaluation datasets. Not included is the additional data for segmentation model training.

#### Ethic approval

The use of the slides from RUMC for the study was approved by the Ethical Committee of the Radboud University Medical Center (2020-7103).

The use of the slides from NKI for the study was approved by the institutional review board of the Netherlands Cancer Institute under number CFMPB737. The use of the slides from SCDC for the study was approved by the Ethic Committee for Clinical Research of the Provinces of Verona and Rovigo under number 25046.

#### Data for segmentation model training

To train the segmentation model, a second multi-centric cohort was assembled, including cases from the NKI, from RUMC collected in previous work [26], and from The Cancer Genome Atlas [27] annotated in [28]. The composition of the training data is described in detail in appendix section S1.2.

### 2.2. Visual TIL scoring

To compare our computational biomarkers with visual TIL-scoring according to the recommendations of the TIL Working Group [6], we set up reader studies for two pathologists to score the NKI TNBC (scored by JS and EM) and the NKI Luminal B cohorts (scored by EM and HMH) using the web-based platforms SlideScore^1^ and CIRRUS Pathology^2^. The pathologists were presented with a web view of a slide, where they could navigate the entire slide and inspect the tissue at different magnifications, but without access to the clinical variables. The pathologists could either give a score from 0 to 100 or mark the slide as unscorable. Only slides scored by both pathologists were used for biomarker development and evaluation, the rest was excluded (see Fig. 2). When multiple slides per patient were available, the slide-level scores were averaged to obtain a single case-level score. We refer to the averaged visual score as vTILs.

### 2.3. Deep learning for tissue segmentation and mitosis detection

As the computer model for tissue segmentation, we chose U-Net [29], a CNN architecture for medical image segmentation. The details of the model and its hyperparameters are described in the appendix S1.3. At test-time, every slide was pre-processed to exclude background and out-of-focus regions using a network that was previously developed and validated [30], therefore only producing a segmentation output for pixels belonging to the biopsy tissue.

The mitosis detection network was used off-the-shelf in this work [24]. In brief, the network predicts the location of mitotic figures across the entire H&E slide. Since the network operates at 40x magnification, to apply the network to the SCDC dataset scanned at 20x, we first upsampled the slides to 40x using bilinear interpolation. Initial visual inspection of the mitoses predictions for slides from the development set showed the presence of false positive detections outside of tumor regions. To address this issue, we combined the mitosis detection with the multi-class segmentation results and only kept mitoses surrounded by tumor at least 20*µm* wide. This distance was determined empirically (see appendix section S1.4 and Fig. S3 for details).

### 2.4. Computational Biomarkers

The segmentation maps and mitosis detections from the deep learning pipeline allow to define biomarkers based on different counts and ratios of the predicted tissues. We designed four morphologically interpretable biomarkers: three related to TILs and one related to mitotic count. The hyper-parameters for the biomarkers, such as values for distances and thresholds, were tuned empirically on the development set to increase pCR prediction performance.

#### Computational TILs

The biomarker cTILs (computational TILs) is aimed to emulate the visual estimation of stromal TILs as proposed by the International TILs Working Group [6]. To this end, the tumor bulk is determined by joining tumor regions within 100*µm* clustering distance and creating an outlining envelope with a 50*µm* margin around them. This is done via the morphological *closing* operation on the predicted tumor mask using a circular kernel with the clustering distance as radius. Then, the tumor mask is *dilated* by the margin distance (see Fig. 3a). In the resulting tumor bulk, lymphocytes and stroma are counted:

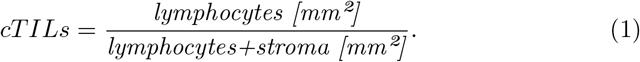

Tumor regions smaller than 0.1mm^2^ were excluded from the tumor bulk formation to account for small wrong tumor predictions.

**Figure 3:**
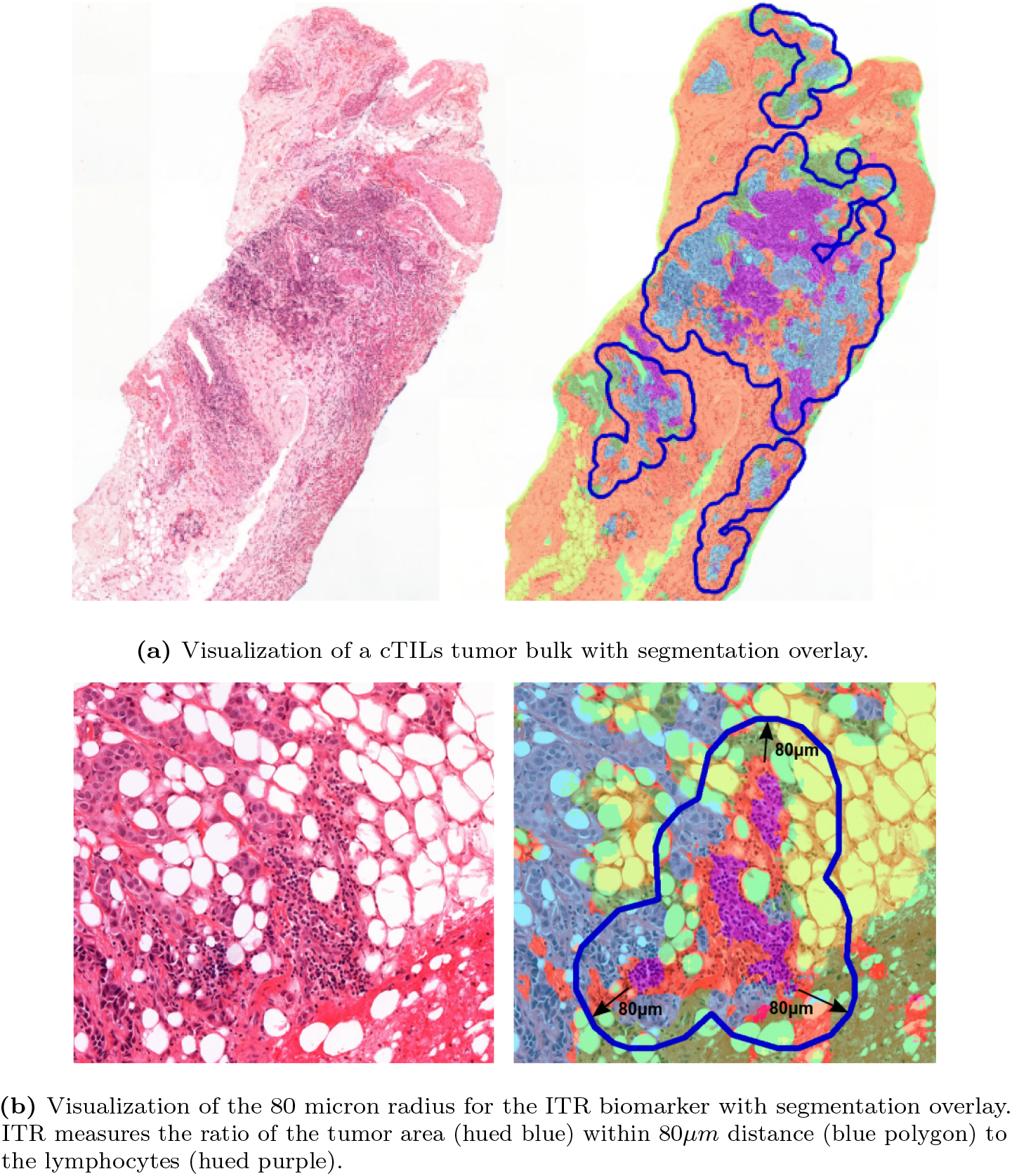
Visualization for cTILs and ITR. In the overlays, tumor is hued blue, stroma orange, lymphocytes purple, necrosis magenta, fatty tissue yellow and the rest green.

#### Lymphocytes to tumor ratio

The biomarker LTR measures the slide-global lymphocytes to tumor ratio (LTR):

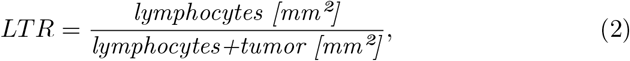

where *lymphocytes* and *tumor* are the predicted area in mm^2^ for the corresponding tissue type from all cores containing tumor predictions.

#### Inflamed tumor ratio

The ’inflamed’ tumor ratio biomarker (ITR) measures the ratio of tumor near lymphocytes to the overall tumor amount:

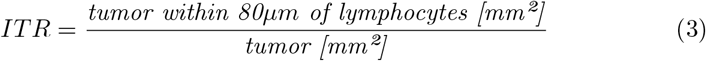

The value for the lymphocyte-tumor ’interaction’ distance of 80*µm* was chosen empirically (an example is shown in Fig.3b; see appendix section S1.6 for details).

#### Mitotic rate

Finally, MTR measures the mitosis to tumor rate:

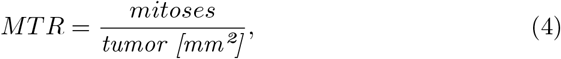

where *mitoses* is the number of detected mitoses inside the segmented tumor regions and *tumor* the amount of predicted tumor in *mm*^2^.

#### Handling multiple biopsies and cores

Usually, a core needle biopsy procedure produces several cores. Only cores containing predicted tumor were considered for the biomarker computation, the rest was excluded. When multiple slides per case were present, the computational biomarkers were computed per case, as if all cores were present on a single slide.

### 2.5. Evaluation and statistical analysis

In order to evaluate the predictive performance of the biomarkers for pCR, we calculated the area under the receiver operating characteristic curve (AUC) and performed multivariable logistic regression, always separately for TNBC and Luminal B. The AUC was computed for the NKI development and evaluation sets and the combined RUMC and SCDC cohorts.

The multivariable logistic regression was performed using the NKI evaluation sets only. The RUMC and SCDC cohorts had too small sample sizes and missing clinical information for proper multivariable analysis. All biomarkers were dichotomized based on their median, except for MTR which was dichotomized as 0 or *>*0, because approximately 60% had a value of 0. The clinical covariates age, grade, T-stage and N-stage were tested as confounding factors. For the MTR biomarker, grade was not tested as confounder, since the mitotic count is part of grading and therefore naturally correlated with grade. For Luminal B, numbers per category were too small in the evaluation set, so no adjusted ORs could be calculated. The covariates were categorized as follows: Age, *<*=50 or *>*50; grade, 2 or 3; T-stage, 1+2 or 3+4; and N-stage, 0 or 1. A covariate was considered a confounder and added to the final multivariable logistic regression model if there was at least 10% change in odds ratio (Exp(B)). The statistical analyses were performed using IBM SPSS Statistical software version 27. The p-values in the multivariable analysis were determined by Wald test per variable.

## 3. Results

### 3.1. Tissue Segmentation

To test the segmentation model, 15 NKI TNBC slides from the development partition were sparsely annotated and verified by a resident pathologist (LT). An example is shown in appendix Fig. S1. The segmentation performance was evaluated by the pixel-wise prediction accuracy. The network segmented 93% of annotated tumor and 84% percent of the annotated lymphocytes correctly; 15% of lymphocytes were wrongly predicted as tumor (see Fig. S2 with the normalized confusion matrix).

The segmentation is the foundation of the biomarkers. An example for the determined tumor bulk necessary to compute cTILs is shown in Fig. 3a. The tumor bulk for this core consists of four regions, from which the lymphocyte and stroma predictions are counted to compute cTILs. An example for the ITR biomarker is shown in Fig. 3b, where the 80*µm*-radius around segmented lymphocytes is marked with dark ovals.

### 3.2. Biomarker evaluation

#### Individual Biomarkers

The AUC results for pCR prediction on all cohorts are listed in Table 1. The ROC-curves for the biomarkers stratified per cancer molecular subtype on the evaluation set are shown in Fig. 4.

**Table 1:** Evaluation results: AUCs, p-values and confidence intervals for predicting pCR for each biomarker and cohort. N and N_pCR_ are the number of cases and responders, respectively. Significant results with p*≤*0.05 are underlined; the best computational biomarker result is in bold font. RUMC+SCDC are the combined RUMC and SCDC datasets.

| Biomarker | AUC | p | 95% CI |
| --- | --- | --- | --- |
| **NKI TNBC development set N=76 (N<sub>pCR</sub>=41)** |  |  |  |
| vTILs | 0.551 | 0.447 | 0.421-0.681 |
| cTILs | 0.479 | 0.759 | 0.344-0.615 |
| LTR | 0.546 | 0.488 | 0.414-0.679 |
| ITR | 0.518 | 0.782 | 0.383-0.654 |
| MTR | 0.494 | 0.929 | 0.362-0.626 |
| **NKI TNBC evaluation set N=66 (N<sub>pCR</sub>=29)** |  |  |  |
| vTILs | 0.681 | <u>0.012</u> | 0.551-0.810 |
| cTILs | **0.766** | <u>0.000</u> | 0.653-0.879 |
| LTR | 0.638 | 0.055 | 0.502-0.775 |
| ITR | 0.712 | <u>0.003</u> | 0.589-0.835 |
| MTR | 0.629 | 0.074 | 0.493-0.765 |
| **RUMC+SCDC TNBC evaluation set N=36 (N<sub>pCR</sub>=15)** |  |  |  |
| cTILs | 0.571 | 0.470 | 0.378-0.765 |
| LTR | 0.568 | 0.490 | 0.372-0.764 |
| ITR | 0.644 | 0.144 | 0.456-0.833 |
| MTR | 0.556 | 0.574 | 0.364-0.747 |
| **NKI Luminal B development set N=276 (N<sub>pCR</sub>=28)** |  |  |  |
| vTILs | 0.739 | <u>0.002</u> | 0.715-0.918 |
| cTILs | 0.651 | <u>0.009</u> | 0.529-0.773 |
| LTR | 0.651 | <u>0.009</u> | 0.531-0.770 |
| ITR | **0.665** | <u>0.004</u> | 0.549-0.782 |
| MTR | 0.652 | <u>0.008</u> | 0.537-0.767 |
| **NKI Luminal B dev. set \| tumor $\geq 8\text{mm}^2$ N=73 (N<sub>pCR</sub>=9)** |  |  |  |
| vTILs | 0.896 | <u>0.000</u> | 0.803-0.989 |
| cTILs | 0.731 | <u>0.026</u> | 0.525-0.937 |
| LTR | **0.748** | <u>0.016</u> | 0.557-0.940 |
| ITR | 0.719 | <u>0.035</u> | 0.514-0.924 |
| MTR | 0.696 | 0.099 | 0.503-0.890 |
| **NKI Luminal B evaluation set N=133 (N<sub>pCR</sub>=9)** |  |  |  |
| vTILs | 0.816 | <u>0.002</u> | 0.715-0.918 |
| cTILs | 0.653 | 0.126 | 0.483-0.823 |
| LTR | 0.516 | 0.872 | 0.320-0.712 |
| ITR | 0.624 | 0.216 | 0.445-0.802 |
| MTR | **0.741** | <u>0.016</u> | 0.570-0.912 |
| **RUMC+SCDC Luminal B evaluation set N=134 (N<sub>pCR</sub>=6)** |  |  |  |
| cTILs | 0.862 | <u>0.003</u> | 0.704-1.000 |
| LTR | **0.888** | <u>0.001</u> | 0.790-0.986 |
| ITR | 0.883 | <u>0.002</u> | 0.718-1.000 |
| MTR | 0.628 | 0.292 | 0.390-0.865 |

**Figure 4:**
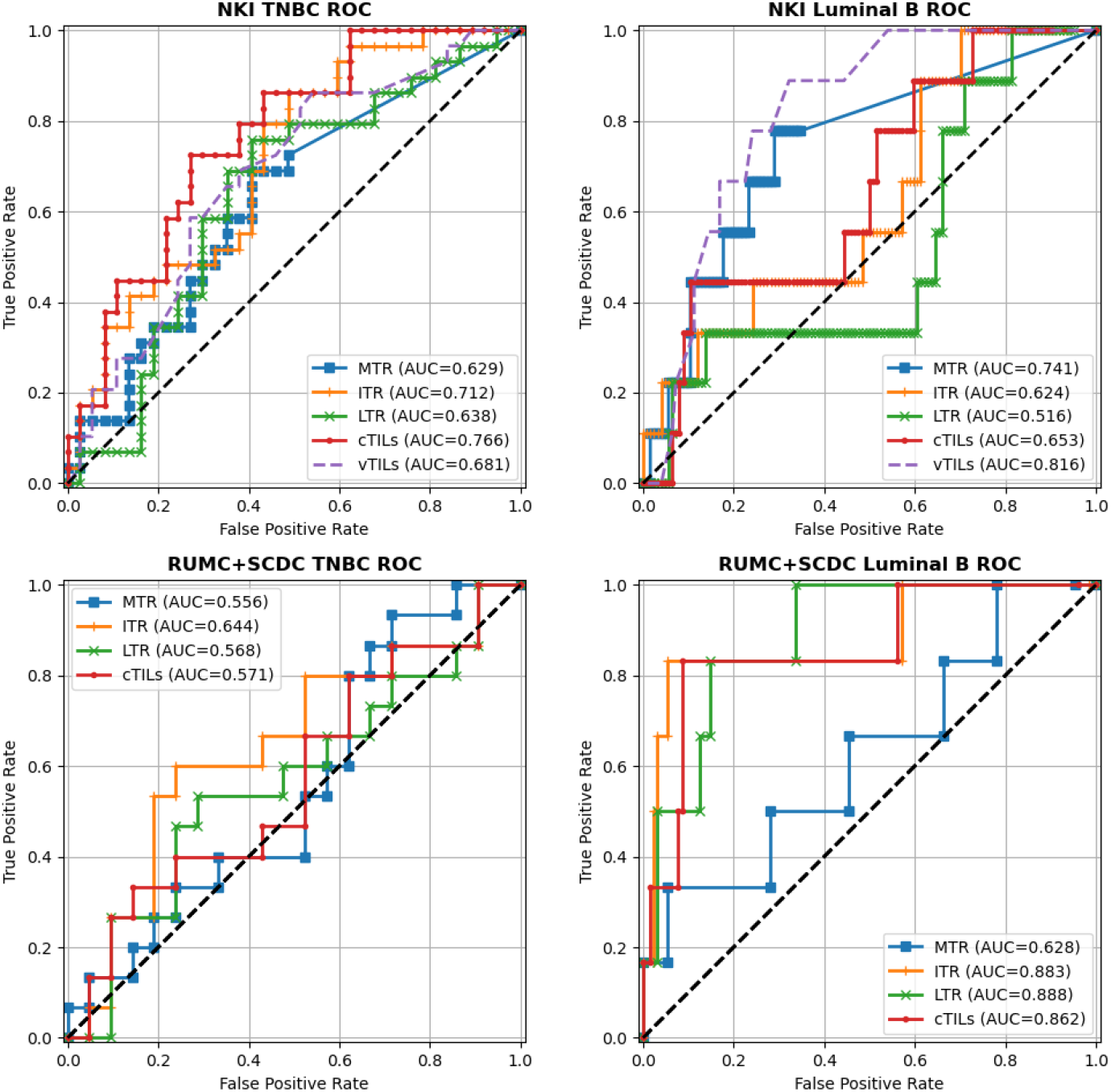
Receiver Operating Characteristic (ROC) curves for predicting pCR on the evaluation sets

For TNBC, all biomarkers have a low performance on the development data, but cTILs and ITR achieve statistically significant results on the NKI evaluation data. The TNBC results for RUMC and SCDC combined (RUMC+SCDC) are not statistically significant.

For Luminal B, ITR exhibits the best performance on the NKI development set and MTR on the evaluation set. On the RUMC+SCDC cases, all biomarkers except MTR reach relatively high scores, however, the number of cases and responders is small. The relatively low performance of MTR on the RUMC+SCDC cases might be connected to the SCDC datasets lower resolution and therefore possibly suboptimal mitoses detection performance.

Each evaluated biomarker has shown statistically significant performance for at least one data subset. LTR achieves no significant result for TNBC, but a relatively high performance on RUMC+SCDC Luminal B. cTILs and ITR perform similarly, with cTILs being slightly better on NKI TNBC. MTR achieves high performance on NKI Luminal B, but its results are not significant for TNBC. Overall, for Luminal B, none of the computational biomarkers reach the performance of the visual TILs-score, while for TNBC cTILs and ITR perform slightly better with only small differences in their performance. A definite comparison, however, is not possible due to the relatively small number of evaluation cases.

#### Influence of the tumor amount on the prediction

The biopsy slides vary in size and (predicted) tumor amount. The TNBC cases from NKI, SCDC and RUMC have median tumor area of 3.7mm², 15.3mm² and 43.6mm², and the Luminal B cases median tumor area of 4.5mm², 8.6mm² and 49.6mm², respectively (combining all slides per case). This raises the question, whether the difference in the available tumor amount has an influence on the predictive performance. To this effect, we investigated the NKI Luminal B development set. First, we verified that the tumor amount itself is not a predictor: The absolute (predicted) tumor-area as biomarker reaches only an AUC of 0.577 with p=0.18. Next, we evaluated the predictive performance on the 72 NKI Luminal B development cases with at least 8mm² of tumor. On this subset, the performance of all biomarkers is increased compared to the full development set (cmp. Table 1). A similar selection from the NKI TNBC development set yielded only 20 cases and no significant biomarker performance (see appendix section S1.7 for more details).

#### Multivariable regression analysis

The statistical results for the logistic multi-variable analysis are shown in appendix Tables S4 and S5. For NKI Luminal B, however, the number of responders was too small to run an adjusted model. For NKI TNBC, the only statistically significant biomarker in the multivariable analysis was LTR (OR 4.57, 95% CI 1.29-16.18; p=0.019); vTILs showed a trend towards significance (OR 2.58, 95% CI 0.85-7.87; p=0.095).

#### Visual TILs

The pathologist’ vTILs biomarker achieves the highest performance on both the NKI Luminal B development and evaluation set and a relatively good performance on the NKI TNBC evaluation set. In comparison, cTILs achieves higher performance on the NKI TNBC evaluation set, but not on Luminal B. The computational biomarker with the highest Spearman correlation with vTILs is cTILs with a correlation of 0.78 for NKI TNBC and 0.57 for NKI Luminal B. The correlation between the individual pathologist vTILs is 0.68 in both cohorts. On the development sets, there is no difference in the performance of the individual pathologist visual scores for NKI TNBC, but for NKI Luminal B the individual pathologist scores achieve AUCs of 0.684 and 0.775. This underlines the subjective manner of TIL-scoring, which can have a strong effect on predictive performance.

## 4. Discussion

### 4.1. Heterogeneity of the biomarkers’ predictive performance

The computational biomarkers exhibit different performances on different subsets of the data with no clear prevalence of a single biomarker. One factor contributing to this is the relative low number of cases due to analyzing them per molecular subtype and center. Another factor might be the available tumor tissue amount. Slides with small tumor regions might not be representative for the whole tumor and therefore do not contain enough information for a reliable prediction. Small tumor amounts might also enhance the effect of minor segmentation errors on the computed biomarkers and lead to deteriorated predictive performance. Evaluation on large, multi-centric external cohorts is required to reliably verify these results. The observed increase of the predictive performance for Luminal B on slides with relative high amounts of segmented tumor indicates that a more reliable pCR prediction is possible, even if only for a subset of cases.

### 4.2. Limitations of the segmentation model

Our trained segmentation model has two limitations that might affect biomarker performance. First, it does not differentiate between invasive and insitu cancer on the assumption that the amount of in-situ tumor co-occuring with invasive tumor would be negligible in core-needle biopsies. Explicitly classifying and excluding in-situ tumor, however, as suggested for TIL-scoring on surgical resections [6], might yield a more fine-grained biomarker performance.

Second, the segmentation model seems sometimes to miss single or sparsely occurring lymphocytes (see. Fig. S1, where single lymphocytes below the red lymphocyte annotation are not segmented). This is probably due to most of the annotated lymphocytes being from clusters of lymphocytes, as these are easier to recognize and annotate. Being able to recognize single lymphocytes would enable more fine-grained biomarkers at the cost of gathering sufficient training data. It is, however, unclear, whether this would improve biomarker performance, since isolated lymphocytes will not contribute substantially to the TILs quantification.

### 4.3. Computational and manual TILs

Scoring of TILs is subject to ’pitfalls’ and bias [9]. This is especially true for biopsies, where less tissue is available and recommendations determined on resections might not apply. Since TILs can be interpreted as the lymphocyte density within the stroma in the tumor-bulk, an important question is how to determine the tumor-bulk when several close-by tumor regions with stroma in-between are present. If such tumor regions are surrounded by lymphocytes and scored individually, the averaged score would be high. However, if the stroma in-between is included, the resulting TILs-score would be low. A large clustering-distance, such as the 750 microns proposed for resections [18], might result in most of the core being included in the tumor bulk. For example, in the slide depicted in Fig. 3a, it would lead to the four tumor-regions being merged into a single encompassing tumor-bulk with a much increased stroma content and only marginally increased lymphocyte content. To avoid such ’under-scoring’ in biopsies, we chose smaller distances and margins in this work. Determining the appropriate settings for biopsies, perhaps also taking the morphological type into consideration, might be required for more stable TIL-scoring, both by pathologists and computationally.

## 5. Conclusion

Predicting pCR is both highly clinically relevant and challenging, as it is currently unknown if the pre-treatment biopsies contain sufficient information for a reliable prediction in clinical routine. Additional factors like the small biopsy sizes and staining artifacts further increase the difficulty level. Nevertheless, we could achieve statistically significant predictive performance with our computational biomarkers while maintaining morphological interpretability. We were able to reach AUCs in the range 0.66-0.88 depending on the cancer subtype and center. These results show that reliable pCR prediction might be possible, even if only for a subset of cases, potentially allowing automated identification of patients at risk of over-treatment. We also showed the predictive value of the mitotic count derived from routine H&E biopsy slides. To the best of our knowledge, this represents a novel contribution of our study to the role of tumor proliferation in the context of neoadjuvant chemotherapy response. Further research will involve validation of the presented techniques in larger cohorts and analyzing additional molecular subtypes such as HER2-positive cases.

## Data Availability

Due to data agreement limitations, the patient data used to develop and validate the biomarkers cannot be shared. The algorithm to compute the biomarkers ITR, LTR and cTILs was made available on the Grand Challenge-platform and can be accessed upon request for research purposes.

https://grand-challenge.org/algorithms/bc-seg-det-rumc

## Acknowledgments

This project was funded by the Dutch Cancer Society (KWF, PROACTING project number 11917), and partly received funding from the European Union’s Horizon 2020 research and innovation programme under grant agreement No 825292 (ExaMode, htttp://www.examode.eu/). We would also like to acknowledge the Core Facility Molecular Pathology and Biobanking at the NKI for technical support and assistance in scanning the H&E-slides. Finally, we would like to thank Dr. Mark Sherman for valuable comments and discussions.

## Data availability

Due to data agreement limitations, the patient data used to develop and validate the biomarkers cannot be shared. The algorithm to compute ITR, LTR and cTILs was made available on the Grand Challenge-platform^3^ and can be accessed upon request for research purposes.

## Contributors

WA performed the experiments, analyzed the results and wrote the manuscript. LM, JH and GB were involved with the data collection. MB was involved in data selection and provided histopathological consultance. EM, JS and HMH scored TILs. YL and AWvdBD performed the statistical analysis. LT supervised manual annotations and checked and assessed the output of AI algorithms (such as the mitoses). FC, JvdL, JW and EHL were involved in supervising the work and experimental design. All authors reviewed the manuscript and agree with its contents.

## Declaration of interests

JvdL was a member of the advisory boards of Philips, the Netherlands and ContextVision, Sweden, and received research funding from Philips, the Netherlands, ContextVision, Sweden, and Sectra, Sweden in the last five years. He is chief scientific officer (CSO) and shareholder of Aiosyn BV, the Netherlands. FC was Chair of the Scientific and Medical Advisory Board of TRIBVN Healthcare, France, and received advisory board fees from TRIBVN Healthcare, France in the last five years. He is shareholder of Aiosyn BV, the Netherlands. MB is medical advisor at Aiosyn BV. All other authors declare no conflict of interest.

## S1. Supplementary Appendix

### S1.1. Clinical information

The Tables S1 and S2 summarize the available clinical information for the cases included in the study.

**Table S1:**
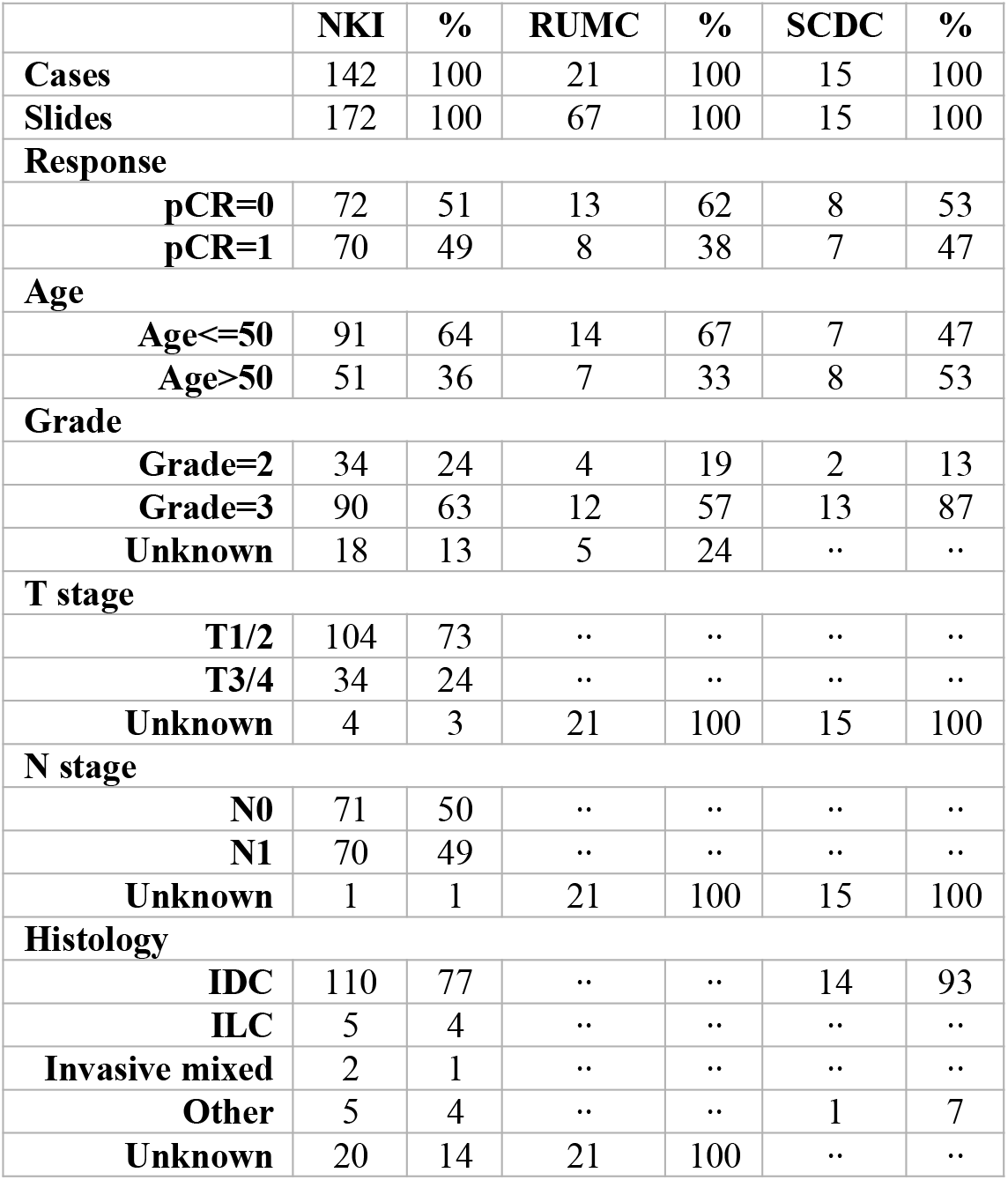
Clinical information for the TNBC cohorts per center (NKI, RUMC and SCDC).

|  | NKI | % | RUMC | % | SCDC | % |
| --- | --- | --- | --- | --- | --- | --- |
| **Cases** | 142 | 100 | 21 | 100 | 15 | 100 |
| **Slides** | 172 | 100 | 67 | 100 | 15 | 100 |
| **Response** |  |  |  |  |  |  |
| **pCR=0** | 72 | 51 | 13 | 62 | 8 | 53 |
| **pCR=1** | 70 | 49 | 8 | 38 | 7 | 47 |
| **Age** |  |  |  |  |  |  |
| **Age≤50** | 91 | 64 | 14 | 67 | 7 | 47 |
| **Age>50** | 51 | 36 | 7 | 33 | 8 | 53 |
| **Grade** |  |  |  |  |  |  |
| **Grade=2** | 34 | 24 | 4 | 19 | 2 | 13 |
| **Grade=3** | 90 | 63 | 12 | 57 | 13 | 87 |
| **Unknown** | 18 | 13 | 5 | 24 | .. | .. |
| **T stage** |  |  |  |  |  |  |
| **T1/2** | 104 | 73 | .. | .. | .. | .. |
| **T3/4** | 34 | 24 | .. | .. | .. | .. |
| **Unknown** | 4 | 3 | 21 | 100 | 15 | 100 |
| **N stage** |  |  |  |  |  |  |
| **N0** | 71 | 50 | .. | .. | .. | .. |
| **N1** | 70 | 49 | .. | .. | .. | .. |
| **Unknown** | 1 | 1 | 21 | 100 | 15 | 100 |
| **Histology** |  |  |  |  |  |  |
| **IDC** | 110 | 77 | .. | .. | 14 | 93 |
| **ILC** | 5 | 4 | .. | .. | .. | .. |
| **Invasive mixed** | 2 | 1 | .. | .. | .. | .. |
| **Other** | 5 | 4 | .. | .. | 1 | 7 |
| **Unknown** | 20 | 14 | 21 | 100 | .. | .. |

**Table S2:** Clinical information for the Luminal B cohorts per center (NKI, RUMC and SCDC).

|  | NKI | % | RUMC | % | SCDC | % |
| --- | --- | --- | --- | --- | --- | --- |
| **Cases** | 409 | 100 | 87 | 100 | 47 | 100 |
| **Slides** | 409 | 100 | 279 | 100 | 47 | 100 |
| **Response** |  |  |  |  |  |  |
| **pcr=0** | 372 | 91 | 83 | 95 | 45 | 96 |
| **pcr=1** | 37 | 9 | 4 | 5 | 2 | 4 |
| **Age** |  |  |  |  |  |  |
| **Age<=50** | 216 | 53 | 55 | 63 | 24 | 51 |
| **Age>50** | 193 | 47 | 32 | 37 | 23 | 49 |
| **Grade** |  |  |  |  |  |  |
| **Grade=2** | 289 | 70 | 52 | 60 | 33 | 70 |
| **Grade=3** | 113 | 28 | 35 | 40 | 14 | 30 |
| **Unknown** | 7 | 2 | .. | .. | .. | .. |
| **T stage** |  |  |  |  |  |  |
| **T1/2** | 288 | 70 | .. | .. | .. | .. |
| **T3/4** | 112 | 28 | .. | .. | .. | .. |
| **Unknown** | 9 | 2 | 87 | 100 | 47 | 100 |
| **N stage** |  |  |  |  |  |  |
| **N0** | 121 | 30 | .. | .. | .. | .. |
| **N1** | 279 | 68 | .. | .. | .. | .. |
| **Unknown** | 9 | 2 | 87 | 100 | 47 | 100 |
| **Histology** |  |  |  |  |  |  |
| **IDC** | 321 | 78 | .. | .. | 43 | 92 |
| **ILC** | 74 | 18 | .. | .. | 3 | 6 |
| **Mixed** | 7 | 2 | .. | .. |  |  |
| **Other** | 4 | 1 | .. | .. | 1 | 2 |
| **Unknown** | 3 | 1 | 87 | 100 | .. | .. |

### S1.2. Data for segmentation model training and testing

First, we included breast biopsies from 89 NKI cases with N=95 slides (82 TNBC, 13 Luminal B, as some cases have multiple slides), and from 15 RUMC cases, where one slide per patient was selected. As a result, a total of N=110 biopsy cases were included. Research assistants, trained and supervised by pathologists, annotated small tissue regions on these slides as tumor, stroma, lymphocytes, necrosis, fatty tissue or rest. Differentiating between tumor, stroma and lymphocytes is essential for TIL-scoring, whereas the other classes were added for a more comprehensive tissue differentiation. An example of two annotations is shown in Fig. S1. The slides were annotated using ASAP^4^. Second, 73 slides were included from a RUMC cohort created in previous work [26], where slides had been sparsely annotated using the same six classes. Finally, we included N=92 slides from the public Breast Cancer Semantic Segmentation dataset (BCSS) [28], containing TNBC resections from TCGA (The Cancer Genome Atlas [27]). These slides contained densely annoated regions of interest (i.e., all pixels in the ROI were labeled) of 18 different tissue types, which we mapped into the six targeted classes for consistency with the rest of the data. Overall, N=275 slides (165 resections, 110 biopsies) were used for model training.

The segmentation model was tested on 15 sparsely annotated NKI TNBC slides. The selection was limited to these slides, due to the heterogeneous nature of TNBC [31], and therefore a more challenging segmentation problem. The test annotations span an area of approximately 12mm² of tumor, 13mm² of stroma, 2.5mm² of necrosis, 2mm² of fat, 0.6mm² of lymphocytes and 0.2mm² of normal glands.

### S1.3. Segmentation Model

In our U-Net implementation we used a depth of four and in the decoder-part bilinear upsampling followed by 2×2 convolutions. We trained the network on patches of size 412×412 px at 20x magnification (0.5*µm/px* spacing) with the patch data-augmentations flip, rotate, scale, color, contrast and noise. The network was trained with learning rate decay starting from learning rate 0.0005 reduced by half after 15 epochs without improvement in validation accuracy. The training was stopped early if there was no improvement after 30 epochs. In each epoch 19.200 patches were sampled with class-wise balanced sampling: each tissue class had the same sampling probability except for fatty tissue, which was sampled five times less frequent.

**Table S3:** Overview of the datasets used for training, validating and testing the segmentation network.

| Source | Type | Training+<br>Validation | Testing |
| --- | --- | --- | --- |
| RUMC [26] | Resections | 73 | .. |
| TCGA [27,28] | Resections | 92 | .. |
| RUMC | Biopsies (neoadj.) | 15 | .. |
| NKI TNBC | Biopsies (neoadj.) | 82 | 15 |
| NKI Luminal B | Biopsies (neoadj.) | 13 | .. |

**Figure S1:**
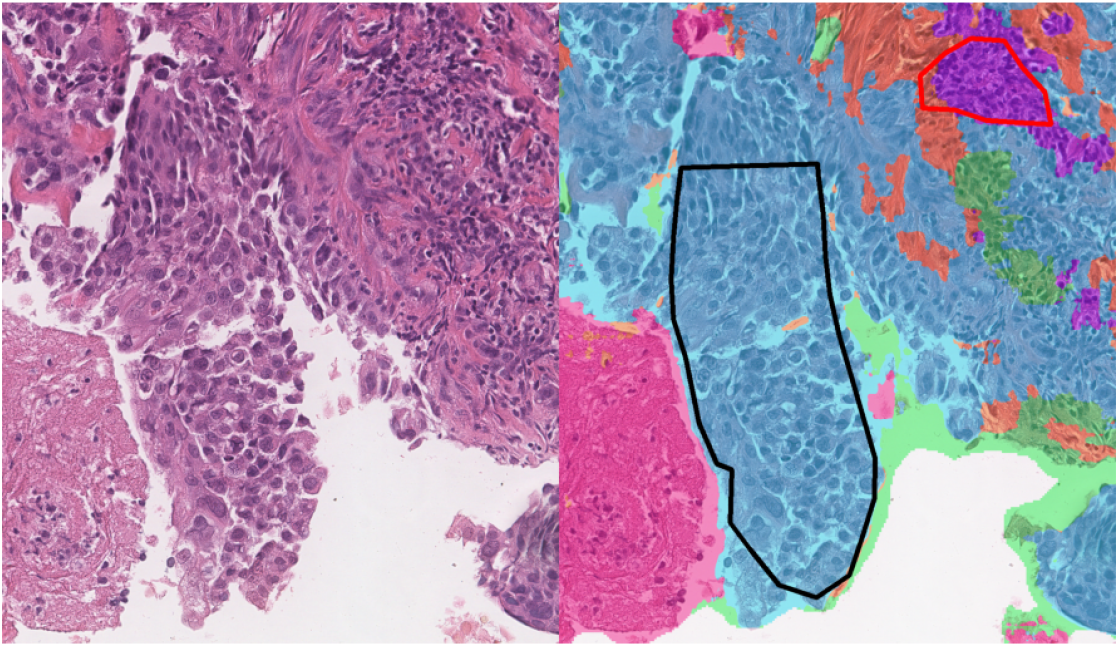
Example from a test slide with segmentation overlay. Predicted tumor is hued blue, necrosis magenta, lymphocytes purple, stroma orange and the rest green. The drawn polygons are the tissue annotations (red: Lymphocytes, black: Tumor).

**Figure S2:**
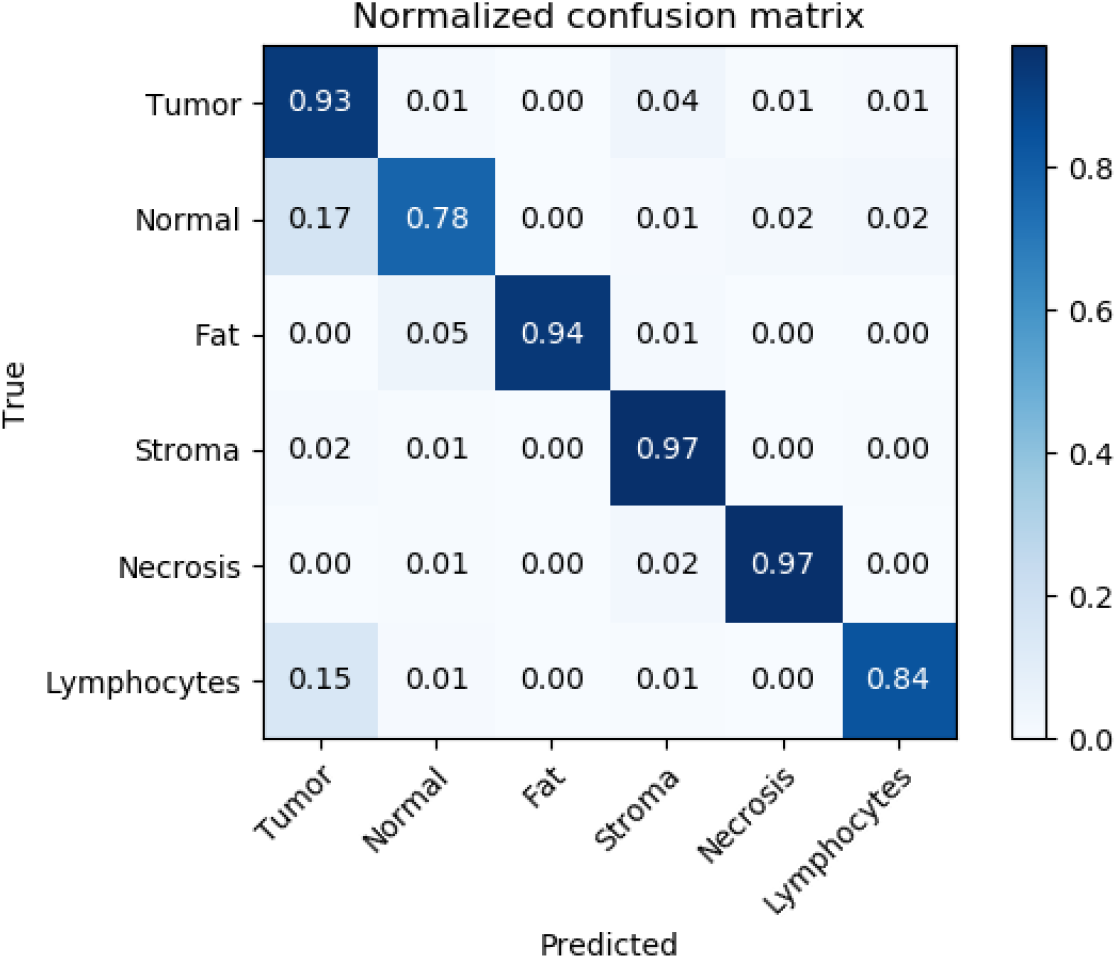
Normalized confusion matrix of the segmentation model for the 15 NKI test slides (computed on the pixel-level). It shows in each cell the ratio of the (true) class on the y-axis predicted as the class on the x-axis.

### S1.4. Mitosis detection

The mitoses predictions on six NKI TNBC slides were checked by a pathologist (LT). Without the tumor-distance filtering, the mitoses recall was 98% with precision 32%, while with filtering the recall was 64% with precision of 60%. Filtering removed around 77% of the detected mitoses on the NKI slides, 55% on the SCDC slides and 59% on the RUMC slides. Fig. S3 shows an example with seven kept and seven removed mitosis detections.

**Figure S3:**
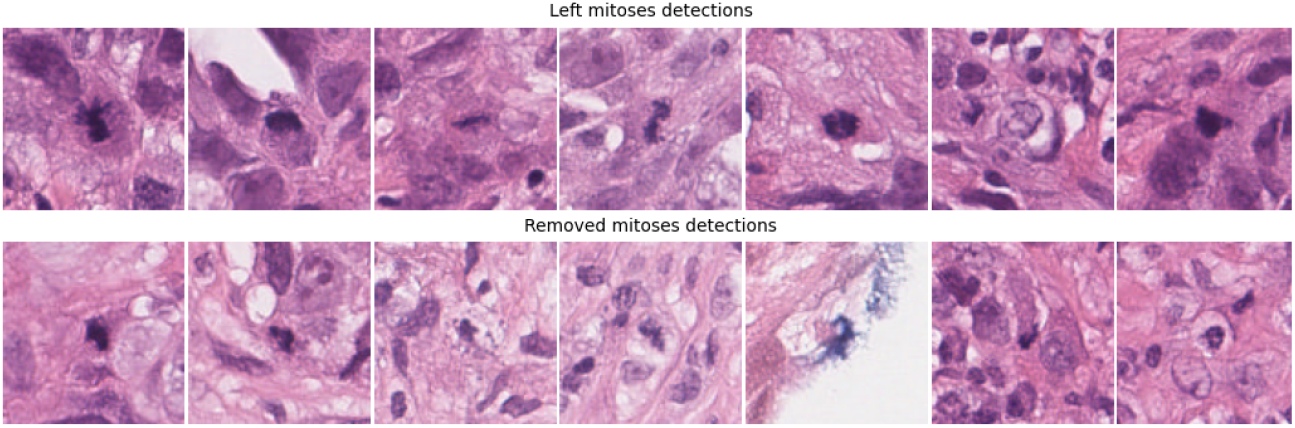
Examples of kept (top) and filtered out (bottom) mitoses detections from a NKI TNBC slide.

### S1.5. Biomarkers

**Table S4:** NKI TNBC multivariable analysis results - unadjusted with only the analysed biomarker and ajdusted together with the clinical variables age, grade, T-Stage and N-Stage. p-values *≤* 0.05 are underlined. *adjusted for grade, ’adjusted for grade and T-stage. nc: no change

| NKI TNBC evaluation set |  |  |  |  |  |  |
| --- | --- | --- | --- | --- | --- | --- |
|  | unadjusted |  |  | adjusted |  |  |
| Biomarker | OR | 95% CI | p-value | OR | 95%CI | p-value |
| **vTILs** | 3.65 | 1.30-10.22 | <u>0.014</u> | 2.58* | 0.85-7.87 | 0.095 |
| **cTILs** | 5.03 | 1.66-15.25 | <u>0.004</u> | 2.37' | 0.69-8.09 | 0.169 |
| **LTR** | 4.61 | 1.57-13.50 | <u>0.005</u> | 4.57' | 1.29-16.18 | <u>0.019</u> |
| **ITR** | 2.79 | 1.02-7.64 | <u>0.046</u> | 1.70' | 0.53-5.48 | 0.373 |
| **MTR** | 2.23 | 0.82-6.08 | 0.116 | nc | nc | nc |

**Table S5:** NKI Luminal B multivariable analysis results - unadjusted with only the analysed biomarker. No adjusted results because of too small number of events.

| NKI Luminal B evaluation set |  |  |  |  |  |  |
| --- | --- | --- | --- | --- | --- | --- |
|  | unadjusted |  |  | adjusted |  |  |
| Biomarker | OR | 95% CI | p-value | OR | 95%CI | p-value |
| **vTILs** | 10.04 | 1.22-82-69 | <u>0.032</u> | .. | .. | .. |
| **cTILs** | 3.18 | 0.64-15.91 | 0.160 | .. | .. | .. |
| **LTR** | 0.43 | 0.10-1.78 | 0.241 | .. | .. | .. |
| **ITR** | 0.93 | 0.24-3.36 | 0.921 | .. | .. | .. |
| **MTR** | 6.59 | 1.31-33.13 | <u>0.022</u> | .. | .. | .. |

### S1.6. Influence of the tumor-lymphocyte distance on ITR performance

The ITR biomarker has a hyper-parameter: the distance between tumor and lymphocytes. This was implemented by dilating the predicted lymphocyte mask and overlapping it with the tumor predictions. We assume that lymphocytes ’close’ to tumor might interact (’attack’) the tumor cells, which is less likely for far off lymphocytes. To our knowledge, however, there are no guidelines on what a suitable distance might be. We therefore evaluated several distances on the development sets - their influence on the predictive performance of ITR for pCR is visualized in Fig. S4. The performance for TNBC is uniformly low, with the best distance around 20 microns. The highest performance for Luminal B is at 100 microns, but the differences between the performances in the 75-125*µm* range are small. The chosen distance of 80 microns for both subtypes retains high predictive performance for the Luminal B development set while maintaining the same level of performance for TNBC.

**Figure S4:**
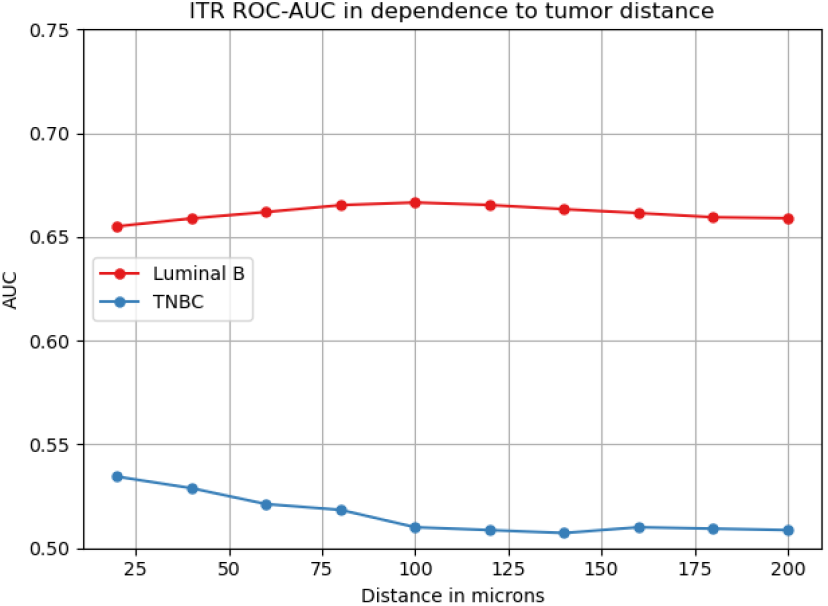
Performance of ITR in dependence to the lymphocyte-tumor distance on the NKI TNBC and Luminal B development sets.

### S1.7. Influence of the tumor amount on the prediction

We also investigated how the segmented tumor amount affects the performance of the biomarkers. Its distribution in the cohorts is visualized in Fig. S5a and its influence is depicted in Fig. S5b and S5c, which show the evaluation of the biomarkers on the NKI development sets partitioned to subsequently exclude cases with less then the given minimal tumor area in mm². This visualization reveals a trend for Luminal B: The pCR prediction improves with increased amount of (predicted) tumor, especially for vTILs.

**Figure S5:**
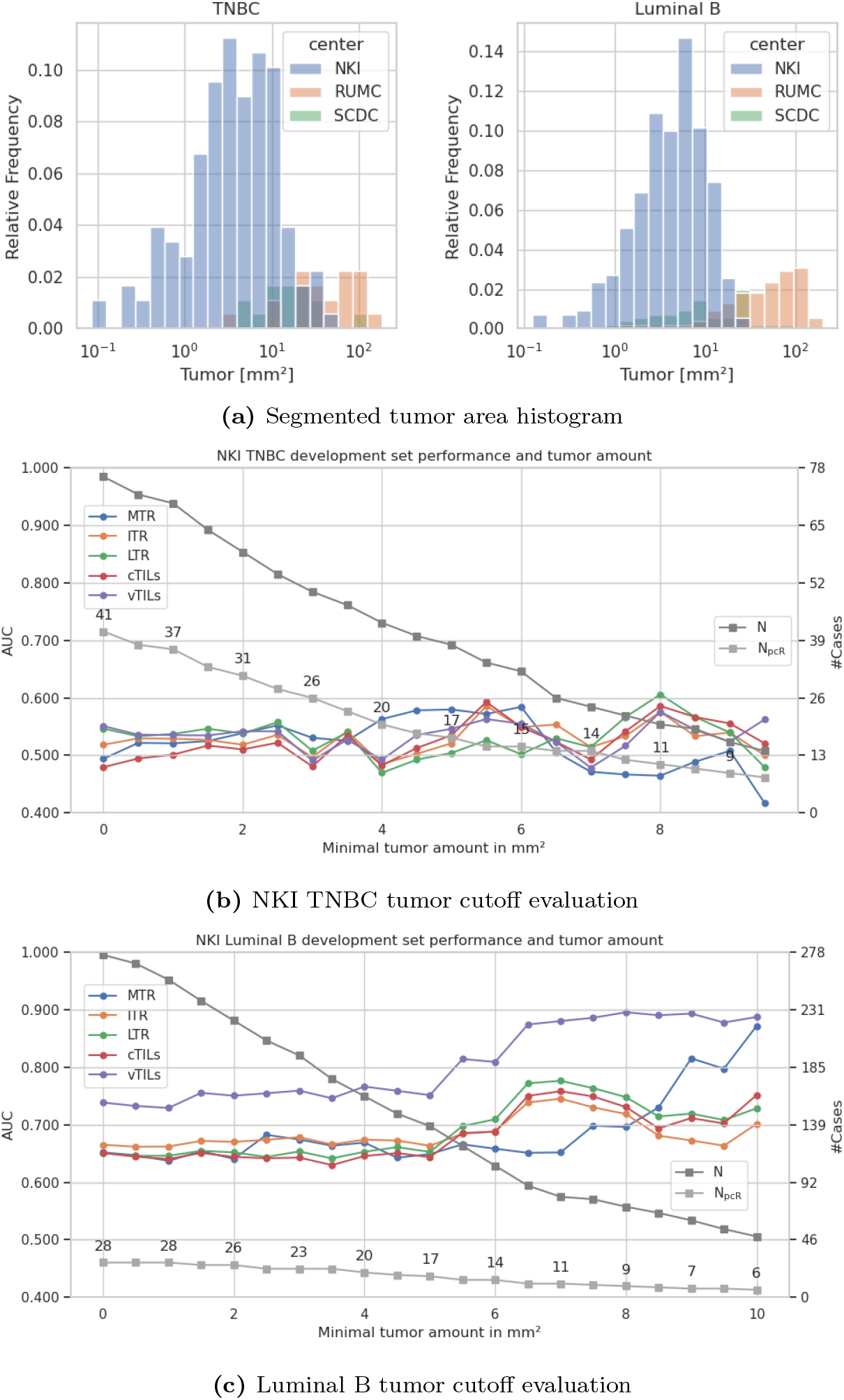
Histogram of the segmented tumor (top) and Biomarker-AUC for pCR evaluated with different (minimal) tumor amount cutoffs on the NKI development set. N and *N*_*pCR*_ are the overall number of cases and responders after the cutoff. The 0mm² cutoff corresponds to the evaluation of all cases. At the 12mm² cutoff only 30 cases with 3 responders are left.

## Footnotes

1 https://www.slidescore.com

2 https://grand-challenge.org/

3 https://grand-challenge.org/algorithms/bc-seg-det-rumc

4 https://github.com/computationalpathologygroup/ASAP

## Notes

### Author Declarations

The use of the slides from the Radboud University Medical Center for the study was approved by the Ethical Committee of the Radboud University Medical Center (2020-7103). The use of the slides from the Netherlands Cancer Institute for the study was approved by the institutional review board of the Netherlands Cancer Institute under number CFMPB737. The use of the slides from the IRCCS Sacro Cuore Don Calabria Hospital for the study was approved by the Ethic Committee for Clinical Research of the Provinces of Verona and Rovigo under number 25046.

### Summary of Updates

Table 1 revised to correct the RUMC+SCDC TNBC evaluation results for the biomarkers cTILs and MTR - the AUC was erroneously inverted (manuscript v2 had still the same error). These changes have no effect on the papers message or conclusion.

